# Guiding principles for accelerating change through health inequities research and practice: A modified Delphi consensus process

**DOI:** 10.1101/2024.04.26.24306421

**Authors:** F. Ahmed, C. Woodhead, A. Hossaini, N. Stanley, L. Ensum, R. Rhead, J. Onwumere, G. Mir, J. Dyer, HSE Collective, S.L. Hatch

## Abstract

Despite a preponderance of evidence, and considerable resources, health & social inequities persist and there is evidence of widening unfair differences in markers of health and care. While power imbalances created by broader structural and economic systems are major influencing factors, reform within health inequities research, policy and health and social care practice is key to both bottom-up and top-down change. We aimed to develop agreement for an iterative set of guiding principles underpinning ways of working for a newly formed Health and Social Equity Collective comprising researchers, community leaders, policymakers, and health and care professionals, seeking to address inequity by identifying and engaging the levers of change within and across institutions. The principles aim to inform a more inclusive and translational knowledge base through research practices, tackling entrenched inequalities in education, training, and capacity-building; and centring communities affected by health inequities through engagement and advocacy. We carried out a modified Delphi consensus process between March and September 2022 with Collective members and networks through online workshops and surveys. Out of 24 consensus statements developed and refined over a workshop and three successive survey rounds, we identified eleven key principles agreed upon by a majority of respondents. Two of these were rated high priority by over 75% of respondents, four by over 60% and five by over 50%. These could be grouped into three main topics detailing ways of working and change needed within: ‘Knowledge and framing of health and social inequities, and incorporation into practice’, ‘Community engagement, involvement and peer research’, and ‘Organisational culture change’. Given the pressing need to address inequities, these principles offer a grounding for future consensus building initiatives which also incorporate a wider diversity of perspectives, and which should be iteratively updated with ongoing learning from health equity initiatives nationally and internationally.

## Introduction

There is an urgent need to accelerate the pace of change in tackling health inequities. For over 20 years, there has been a focus on identifying and tackling health and social inequities in research and policymaking in the UK. However, such inequities persist and are widening [1–3], exacerbated by the COVID-19 pandemic [4]. Translation of the vast industry of health inequalities research into practice is limited at best, self-serving at worst [5,6]. In research and policymaking, current ways of working and of training the next generation across disciplines and sectors contribute to this stagnation, being inherently fraught with inequities and fragmented approaches [7–9].

Across research, education, policy, public health, and healthcare sectors, there has been an overwhelming focus on inequalities (i.e., unequal distribution of health and social outcomes), despite an amassing of evidence highlighting a lack of attention to the existing and emerging inequities (i.e., injustices) experienced by groups within and across institutions and systems [10–12]. Notably, the focus on social determinants and on individualised behaviour change approaches have paid little attention to the main systematic and institutionalised drivers and processes generating, maintaining, legitimising, and perpetuating inequities, such as racism, xenophobia and all forms of discrimination [13–15]. These are experienced, anticipated and witnessed across the life course in different domains, including health, education and research institutions [16]. Moreover, while much deficit-based work has examined the experience of marginalisation on health and well-being, there has been very little acknowledgment of the impact of privilege in creating, maintaining and perpetuating health inequalities [17]. Similarly, a lack of consideration of the impact of norms, behaviours, laws, processes enacted inter/intra-personally, organisationally and structurally which maintain advantage among individuals and groups benefitting from the status quo persists [18,19].

By actively disrupting predominant power structures, ‘empowering’ those affected by health inequities through participation and involvement in research and decision-making is increasingly viewed as a key mechanism of change [20,21]. However, approaches such as ‘coproduction’ and ‘community engagement’ in research and practice often fail to achieve the empowerment required for systems change [21–23]. There is a need for more equitable and adequately resourced approaches to engagement which facilitates capacity and relationship building between organisations and community groups affected by health inequalities; spaces for dialogue which explicitly and sensitively attend to power differentials; institutional culture shifts which valorise community voice; and, transparent acknowledgement of historic and contemporary abuses of power which inhibit trust [21,23,24]. The coproduced Patient and Carer Race Equality Framework (PCREF), launched as the NHS’ first anti-racism framework, is one example of a nationwide participatory approach to tackle mental health inequities, which has community engagement at its core.

Health inequities are also shaped by wider structural social and economic forces; however, processes and practices in research, education, health and social care form part of the system maintaining health inequities in place. There is a need to develop shared agreement on best practice principles for how to initiate alternative, sustainable, and interdisciplinary ways of working and ideological shifts to help tackle the structures which perpetuate inequities in what gets researched, whose voices are heard, how training and career pipelines are navigated and what evidence is valued [8]. As we note, structural inequity relies on political, economic and ideological forces which outmass the sectors we represent. Thus, we aim to change what we can within our domain while searching for points where we influence broader social currents.

As a foundation for facilitating such changes in our work we formed the Health and Social Equity Collective (www.hsecollective.org) in November 2021, a group of leading interdisciplinary researchers in the field of health equity, community advocates, policy makers, health and social care professionals and voluntary and community sector organisations with the goal of accelerating the pace of change in tackling health inequalities. The composition of the Collective supported a focus on a broad range of socially excluded groups (e.g., migrant, racialised, religious minority, sexual and gender minority groups, people with mental ill health, learning disabilities and disabled people), as well as research approaches. The first phase of the HSE Collective was an extensive co-design process, during which we developed our priorities, ways of working and public facing resources. As part of the co-design phase, we conducted a modified Delphi exercise which aimed to incorporate the perspectives of people affected by, and working to address, health inequities to build consensus about how best to change the way we work and collaborate to achieve those broader aims across three domains: 1) research, 2) training and capacity building, and 3) engagement and advocacy.

## Methods

Ethical approval was obtained 15^th^ February 2022 from the King’s College London PNM Research Ethics Panel, reference: RS/DP-21/22-28451.

### Convening the collaborators

We undertook a modified online Delphi consensus building process [25] in line with previous approaches within health and social care research [26–28]. A diverse group of collaborators was formed in November 2021 (HSE Collective) with the first workshop being held a month later. Ethics were granted on February 15th, 2022, and the first round of consensus building began on March 1st, 2022. This was a way of integrating geographically disparate expert perspectives (including lived experience) on key areas for change in practice across the research, training and capacity building, and engagement and advocacy domains through successive rounds of prioritisation, rating and iterative refinement of a series of statements. An outline of the approach is detailed in Table 1 below, following that conducted by Mir et al. [27].

**Table 1.**
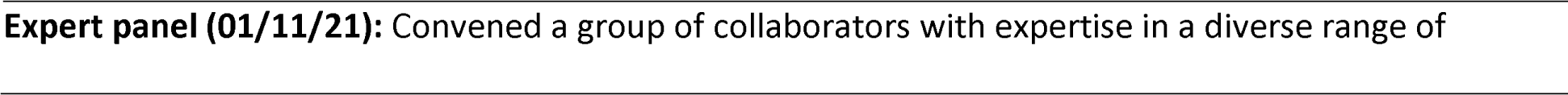
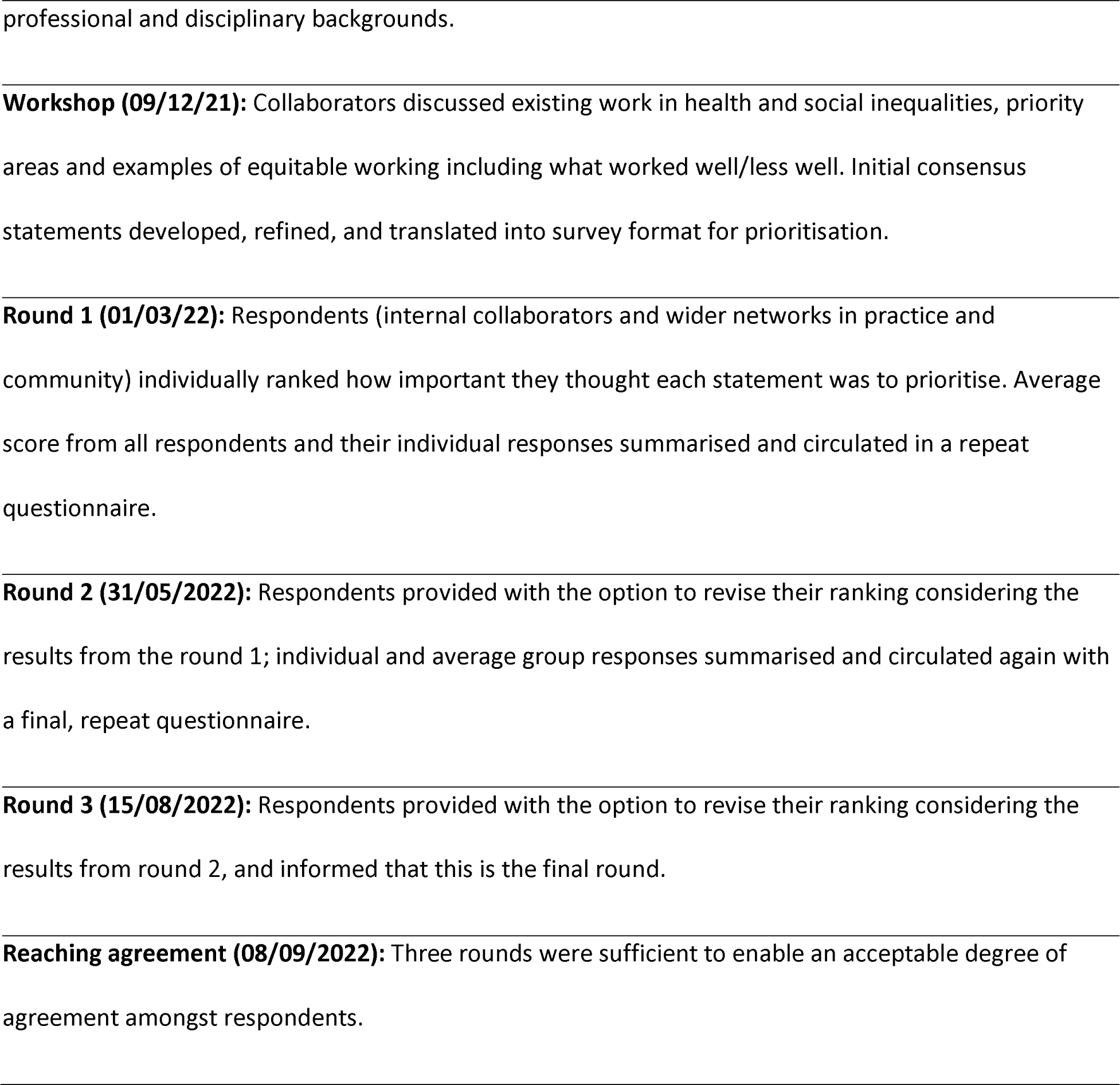
Main steps in the Modified Delphi exercise.

Collaborators within the Collective working across academia, policy, health and social care, arts and culture, and community and voluntary organisations in the UK and internationally were identified through professional networks (growing to 76 members). They were invited to attend an online workshop, which consisted of small-group discussions in breakout rooms to discuss: their work relevant to health inequities, priorities for future research and practice, how they have approached the issue of equity and how it is embedded in partnerships and any learning from prior work or experience, e.g., in terms of attending to inherent power dynamics. The key aim of this initial priority-setting workshop was to obtain content that would inform the consensus statements for the Delphi consensus process.

### Developing the consensus statements and initial survey

We conducted a rapid data gathering exercise involving an online search of websites and resources from other health equity-based initiatives and comparator organisations nationally and internationally. This information, and content from the initial workshop relevant to the three HSE Collective domains (research, education and capacity building, and engagement and advocacy) was collated and thematically organised by topic. Based on this, we next drafted initial consensus statements (n=23) arranged under six themes: 1) Intersectional training and education; 2) Challenging language and narratives; 3) Capacity building; 4) Equitable career pathways; 5) Shifting power; 6) Data accessibility and transparency. Statements were reviewed to ensure clarity, removing duplicate concepts and/or refining similar concepts into a single statement, removing unnecessary jargon and assessing interpretability and accessibility.

Next, we developed an online survey which included the initial consensus statements, asking respondents to score statements in order of priority against a 4-point Likert scale ranging from ‘1 = not a priority: we should not pursue this ‘, ‘2 = low priority: we should pursue this when possible and when other issues are addressed’, ‘3 = medium priority: we should pursue this after a high priority is addressed’ and ‘4 = high priority: we should pursue this now’. Each statement also provided the option to make additional free-text comments relating to the statement’s clarity, ideas for how to amend or change the statement, or other views on the statement. Questions about socio-demographic characteristics (age group, race, ethnicity, migration status, employment status and sector, sexual orientation, gender, any religious identification, education level, long-term physical, sensory or mental impairment, and caring responsibilities) were included to appraise whose voices were being represented among respondents, and to facilitate targeted dissemination to capture a wider variety of perspectives. Given the inclusion of international collaborators, we refined socio-demographic category terminology to reflect contextual differences.

The survey included an embedded information sheet detailing information about data protection, confidentiality and withdrawal procedures. Before starting the survey, respondents were asked to complete a consent form, which also asked for their e-mail address and consent to be e-mailed further survey rounds. Links to the survey and information were sent to existing collaborators, who were invited to complete it themselves and to share information about the survey and the link with their wider networks. We particularly asked if they could share with people or groups with lived experience of health and social inequities and other forms of marginalisation (e.g., via research/staff networks, advisory groups, personal contacts). Each survey round was open for a one-month period.

### Data analysis and further rounds

Using the in-built survey reporting function, aggregate descriptive statistics (i.e., number and percentage) were captured across socio-demographic items. Next, open-text comments were extracted and reviewed, revising statements accordingly by rewording, re-ordering, grouping together and removing statements, leaving 21 revised statements.

Statements were circulated again in the same survey format via e-mail for round 2 of the Delphi exercise. This time, statements were not separated into themes to enable respondents to think about them independently as standalone items. Each respondent who participated in round 1 received their original score in addition to the average scores for each statement (i.e., the percentage of respondents who scored the statement not a priority, low priority, medium priority and high priority). This provided respondents with the option to re-evaluate their position based on the wider feedback.

Round 2 statements and socio-demographic data were analysed and refined in the same way as for round 1. At this stage, the number of statements were increased to 24, based on the suggestions and feedback received in the open-text comments. The round 3 survey was developed and disseminated in the same way, though statements were presented as one list and rearranged so that respondents would view and appraise them in a different order. Using similar methods to prior approaches (e.g., Mir et al., 2012), consensus was initially defined as >75% of respondents ranking the statement as either medium or high priority, and the consensus statements were based on the results of round 3 (the final round of the Delphi exercise), with some minor amendments to wording of the final prioritised statements where open text comments asked for greater clarity.

## Results

A total of 21 collaborators attended the initial workshop including representatives from policy (n=1), arts and culture (n=4), and voluntary and community sectors (n=4), health and social care (n=3), and academic researchers (n=9). After developing and disseminating the Delphi survey, a total of 40 people responded to the round 1 survey. The majority of respondents were aged 25-54 years (83%) and identified as female (80%), just under half were from minoritised racial or ethnic groups while over a fifth identified as a sexual minority. Those migrant to the country they currently reside, those identified with any religion and those with any longstanding mental, physical or sensory impairment each made up around a third of respondents. Most were employed and had degree-level education or higher. Just under half were health and social care professionals, with academic researchers comprising over a third of respondents. Subsequent response rates were n = 28 in round 2, and n = 28 for round 3. Those responding to rounds 2 and 3 tended to be older and included a greater proportion of academic researchers than round 1. Round 3 statements and scores are outlined in Table 2, while the final consensus statements following minor amendments to wording are summarised by topic in Table 3 below.

**Table 2.** Summary of round 3 statements and scores.

| **Consensus statements** | ***n*** | **High**<br>**priority**<br>**score**<br>**(%)** | **Medium**<br>**/ high**<br>**priority**<br>**score**<br>**(%)** |
| --- | --- | --- | --- |
| There is a need to advocate for the integration of knowledge about social and structural determinants of health and social inequities (racism, homophobia, transphobia, sexism, ableism, ageism, etc.) in academic and professional training and curricula. | 28 | 82 | 100 |
| When explaining health inequities, problems should be traced to social and structural systems of oppression rather than located within affected individuals and communities.<br>*\* Social and structural oppression refers to a combination of prejudice and institutional power (manifested in norms, policies, processes, laws etc.) that regularly and consistently discriminates against some groups and benefits other groups)* | 28 | 82 | 100 |
| Health and social care staff should consider people's intersecting social identities/positions when diagnosing and caring for people, while ensuring that presumptions and stereotypes are not reinforced. | 28 | 64 | 100 |
| To foster healthier and more equitable ways of working, the capacity (e.g., time, energy, accessibility) needs of socially excluded groups | 28 | 61 | 100 |
| should be considered prior to consultation and it is important to consult with them before designing their participation or involvement. |  |  |  |
| To ensure fair career progression for socially excluded groups, culture change initiatives are required to improve recruitment, retention and promotion and provide better support in work and learning environments that are traditionally exclusionary/hostile. | 28 | 61 | 93 |
| There is need for higher standards and expectations from funders for inclusive practice within the processes outlined in the above statements, and an acknowledgment that this level of commitment and resources exceeds those linked to traditional/conventional approaches. | 28 | 61 | 93 |
| There is a need to develop new, and evaluate and share existing tools, resources and methods which help empower communities and amplify the effectiveness of community engagement.<br>*\*Community engagement means involving communities in decision-making and in the planning, design, governance and delivery of projects and services.* | 28 | 57 | 100 |
| There is a need to embed monitoring and mechanisms of accountability for senior leadership/management to ensure that Equality, Diversity & Inclusion initiatives result in positive change. | 28 | 57 | 100 |
| Where possible health inequities research should involve and support peer researchers (people with lived experience of the issues | 28 | 57 | 96 |
| of interest direct and conduct the research) to help ensure it is meaningful and accessible to groups most affected. |  |  |  |
| There is a need to provide stakeholders (e.g., community members/organisations, researchers, policy makers, service designers) with training, resources and skills to recognise existing expertise and skills amongst marginalised communities and to challenge hierarchies within decision-making processes. | 28 | 57 | 93 |
| It is important to understand and explicitly state how narratives, norms, processes and practices systematically benefit advantaged groups, including by focusing health inequities research on privileged as well marginalised groups. | 28 | 54 | 96 |
| Staff require appropriate support (e.g., with protected time, resource/tools, encouragement & direction) to help them ensure that their policies, practices and decisions are fair, meet the needs of, and are not inadvertently discriminating against any group(s). | 28 | 50 | 96 |
| To tackle community hesitancy about collaborative research, transparency about data ownership (especially data sovereignty and governance), intellectual property, ethical standards, compensation, accountability mechanisms and the intentions of each stakeholder should be presented from the outset. | 28 | 50 | 93 |
| There is a need to use a range of types of data and to adapt data presented to specific audiences (e.g., academic, health and social care, and community/voluntary sector) to be most impactful. | 28 | 50 | 96 |
| There is a need to amplify and acknowledge resistance, agency and everyday thriving as opposed to resilience and deficit-based perspectives when writing about and working with marginalised communities. | 28 | 50 | 89 |
| Methods for enabling community participation and partnership working should be standard elements of research, policy and practitioner training curricula. | 28 | 46 | 89 |
| To ensure fair career progression for socially excluded groups, there is a need to develop alternative career pipelines such as apprenticeships, paid internships, professional services secondments and placements, and flexible entry requirements that are equitable, inclusive and shaped around the needs of communities. | 28 | 46 | 96 |
| There is a need to achieve consensus on a framework underpinning the incorporation of community engagement and participatory approaches into research, policy, and service design.<br>*\*This includes issues such as equitable working, trust-building, recognising capacity issues such as fatigue, engaging in reciprocity and sustainability of relations and initiating conversations around power and privilege.* | 28 | 43 | 89 |
| There is a need for transparent evaluation of the implementation, monitoring and impact of Equality, Diversity and Inclusion (ED&I) initiatives within health and social care, education, research, funding practices and policymaking. | 28 | 43 | 93 |
| To highlight the extent of inequity affecting marginalised groups, there is a need to mandate and/or incentivise collection of standardised & monitor quality of data on social demographics and where possible use finer-grain, more disaggregated categories relevant to local circumstances. | 28 | 43 | 93 |
| Creative productions such as photography, drama, video, and public events are an effective way to document and disseminate evidence on health and social inequities. | 28 | 36 | 89 |
| To ensure fair career progression, health and social care employers and research institutions should adopt explicit proactive measures/actions (e.g., which are anti-racism) that are subject to transparent and accountable evaluation and monitoring with clear communication on the consequences of falling short on these standards. | 28 | 32 | 93 |
| Creative mediums should complement traditional scientific methods in research about health and social inequity. | 28 | 32 | 86 |
| Opportunities for sharing knowledge and examples of good practice in reducing health inequities should be embedded within and across sectors, including those which are not directly related to healthcare. | 28 | 29 | 100 |

**Table 3.**
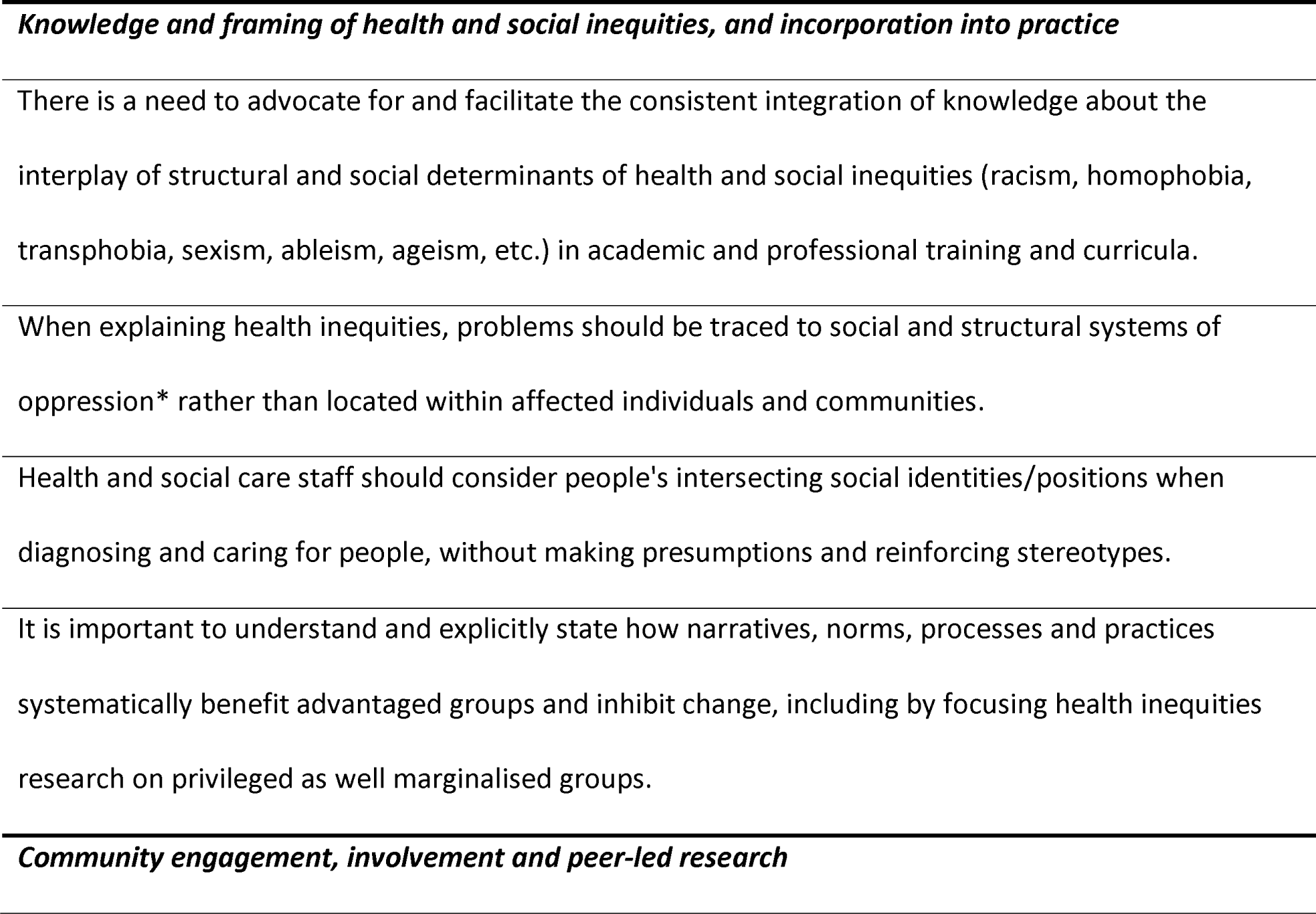
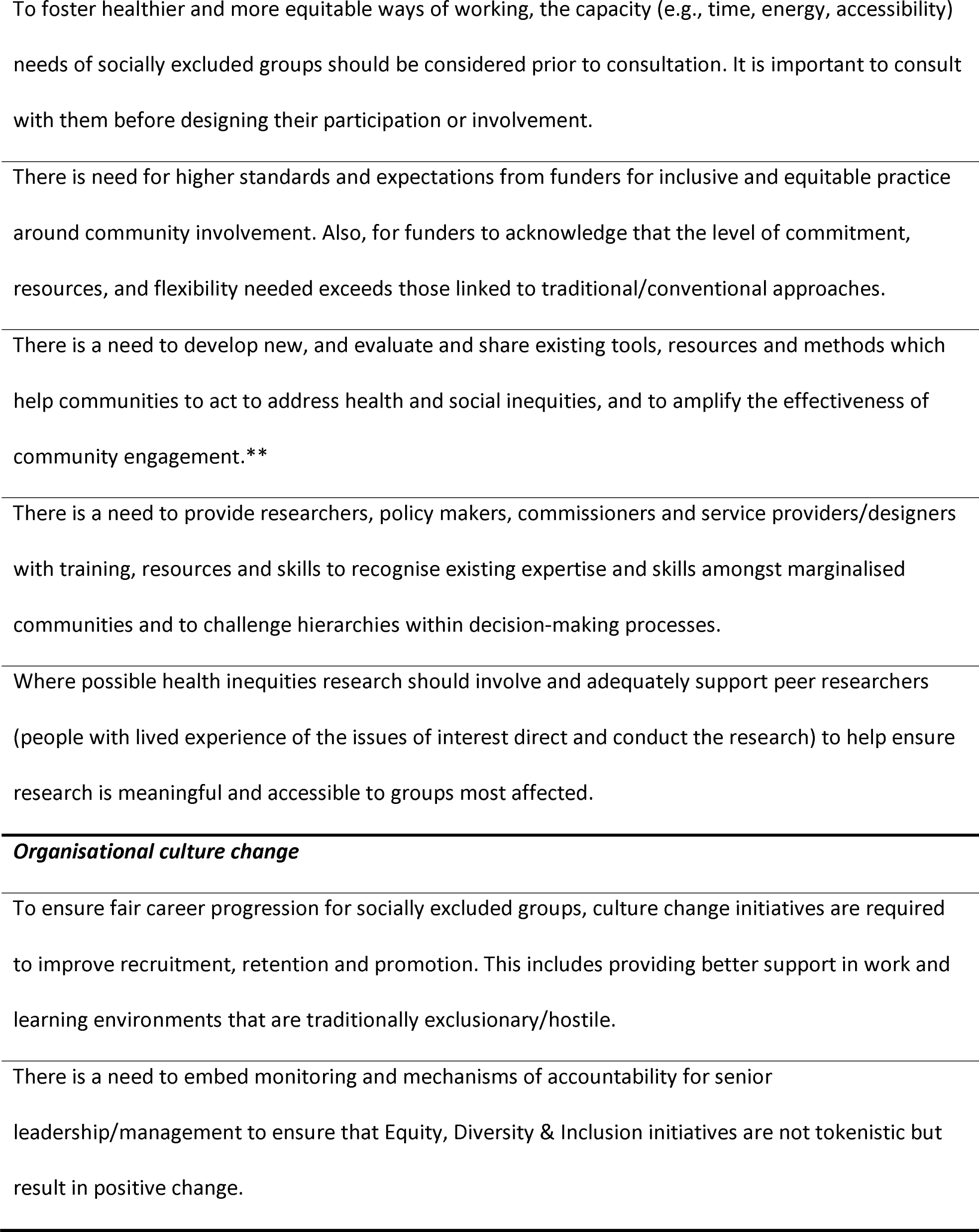
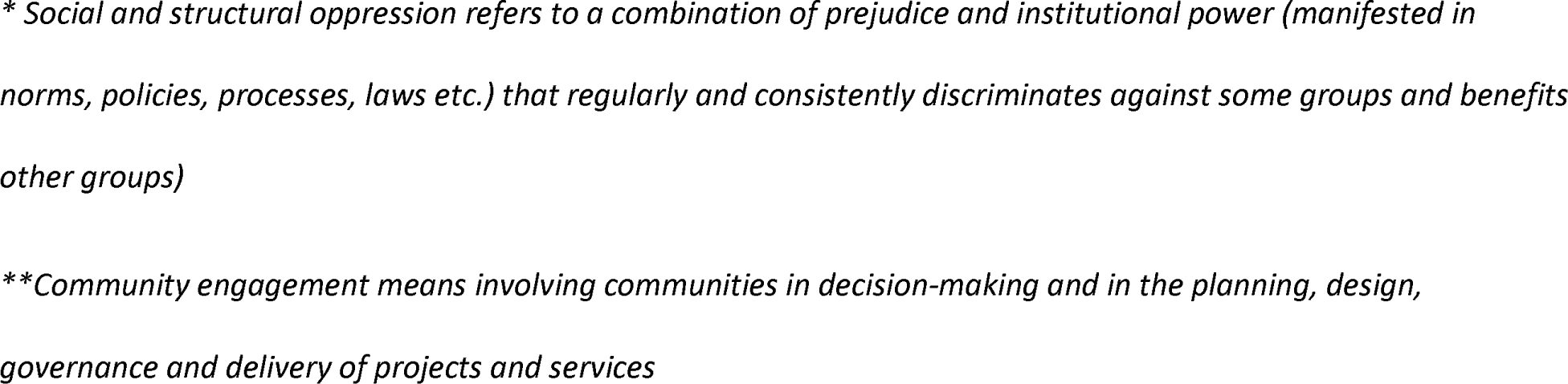
Final consensus statements, grouped by topic.

As shown in Table 2, all 24 statements met consensus criteria (>75% scoring as either medium or high priority). To allow for further prioritisation and to distinguish between these, we therefore examined the proportions scoring each item as a “high” priority only. According to this criterion, only two statements met the threshold. Given the more stringent approach, and in line with other published approaches [29], we expanded the threshold to include those items for which >60% scored as high priority (corresponding to six statements), and those for which over half of respondents scored the statement as a high priority (a further five statements). By presenting statements against each of these tiers (>75%, >60% and >50% scored as high priority), we therefore indicate those statements for which the majority of participants report as high priority.

### >75% scored as high priority (n=2)

Two statements in this category referred to increasing knowledge about social and structural determinants of health in professional training and academic curricula, and to ensure that the causes of social inequities are understood as resulting from patterns of social and structural power rather than being located within the affected groups themselves. Open text comments emphasised the need for help in facilitating such learning, to ensure that it is continually reinforced and not part of mandatory one-off tick box training. These statements were viewed as overarching and facilitative of other practice-related consensus statements (e.g., feeding into how health and social care professionals consider people’s intersecting identities when caring for them). Understanding the structural forces that shape and pattern health and social inequities was perceived to be essential to move away from individualised approaches to care delivery, service design and public health planning, and to mitigate the harm from poorly framed health inequities research which problematises those affected.

### >60% scored as high priority (n=4)

Linked to the first two statements, the next priority referred to integrating understanding about structural determinants and social power linked to people’s intersecting identities within their approach to care, while avoiding making assumptions or stereotyping. This approach was seen as important to correcting the assumptions which underpin many decisions and the resources available for appropriate treatment, to mitigate the risk of people refusing or opting out of care that is insensitive to different aspects of their identities, and to support a focus on challenging inequities inherent to current ways of working.

While the first three emphasised health and care practices, one statement in this category referred to the need to attend to norms and cultures shaping workforce and career progression inequities within institutions/organisations responsible for research and care delivery. The focus on organisational culture and norms which are exclusionary to people from marginalised groups distinguishes this statement from a related statement on career progression given lower priority, relating to the need for transparent evaluation and monitoring of proactive measures mitigating inequity (e.g., which are anti-racism), with accountability for falling short. Open text comments indicated that the latter was perceived as insufficient alone, lack of clarity about what such measures should be, and that initiatives should be intersectional rather than focused on single identities.

The two remaining statements focussed on meaningful and equitable involvement of people with lived experience of social inequities in research, service and policy design, oversight and delivery. They highlight the importance of involvement that is adequately planned to be considerate of their needs and adequately resourced in terms of time and funding - with flexibility to account for people’s lived reality rather than adhering to strict timescales. Open text comments emphasised the importance of allowing sufficient time for involvement and participation, and that without consideration of people’s capacity, time and energy, and adequate renumeration, any community-led initiatives will be seen as serving the institutions/organisations rather than in their best interests, and thus extractive and counterproductive.

### >50% scored as high priority (n=5)

Three of the five statements for which over 50% of respondents scored as high priority also related to involvement of people with lived experience and expertise in research, service and policy design, oversight and delivery. Specifically, in developing and sharing tools to support community engagement and community-led initiatives and working equitably with sufficiently supported and resourced peer researchers adequately supported to take part. Further, one statement emphasised the importance of training and support for those involved in research and decision making to recognise existing expertise and skills amongst marginalised communities and to challenge hierarchies within decision-making processes. Several open text comments indicated that while such efforts are important, they challenged the idea of academic-led community engagement and empowerment. They emphasised the facilitative role of the Collective, such that involvement is community-led, while also facilitating health and care services and organisations to meaningfully involve people with lived experience and expertise themselves. Comments also emphasised the need for tools or resources to support trauma-informed engagement practice and to proactively form more equitable partnerships which meaningfully share power rather than recreate existing power dynamics. Regarding peer-researchers, other considerations emphasised the need for flexibility in relation to their time and capacity as well as sharing credit for work undertaken to avoid tokenistic and exploitative involvement.

One statement in this category overlapped with statements in the >75% category, emphasising awareness and acknowledgement of how narratives, norms, processes and practices systematically benefit advantaged groups. Open text comments suggested that this statement should be considered alongside other related statements in an overall approach which shifts perspectives towards the structural and systemic factors creating and maintaining patterns of inequities. This would support moves away from deficit and damage-centred approaches and help identify points for change within processes and practices.

Finally, the statement referring to the need to embed monitoring and mechanisms of accountability for senior leadership/management to ensure that Equity, Diversity & Inclusion (ED&I) initiatives result in positive change, was endorsed by over 50% of respondents. Open text comments supported this statement but warned against tick box and tokenistic approaches (e.g., online one-off trainings or poorly designed Equity Impact Assessments) that were overly onerous for organisations. One comment suggested that the impact of ED&I initiatives would be dependent upon accountability frameworks and the extent to which they were embedded into inspectorates with capacity to sanction (e.g., Ofsted, Care Quality Commission). Another emphasised the importance of considering how social class intersects with other forces of oppression and privilege to shape people’s experiences to avoid misleading representation of progress made (e.g., where greater equity for women was only experienced among women from affluent backgrounds).

Taking these comments into account with minor refinements to wording, and ordering them according to topic, the final 11 consensus statements are provided in Table 3.

## Discussion

We gained majority agreement around eleven statements pertaining to high priority ways of working within research, education and policy/advocacy that an expert panel identified as priorities for meaningful change in health inequities. These could be grouped into three main topics: *‘Knowledge and framing of health and social inequities, and incorporation into practice’, ‘Community engagement, involvement and peer research’, and ‘Organisational culture change’*.

### Knowledge and framing of health and social inequities

The need to incorporate information about social determinants within health and care professional training and development has been established [30] with evidence for improved knowledge and confidence in addressing social determinants [31]. However, the extent to which this has been implemented is unclear, especially beyond the US context and beyond medical training [31]. Moreover, it is unclear whether, or the extent to which, such training involves reflexive awareness about how individual and organisational power operates to influence health and care inequities, with some notable exceptions [32,33]. Findings suggest that such learning should extend beyond practitioners to include those training in a broad range of health-related disciplines (e.g., epidemiology, statistics, public health). In research, there has been increasing awareness of the need to move away from damage-centred and deficit perspectives (i.e., focusing just on adversity and it’s adverse impact on affected groups) in research on health and social inequities [34], and towards approaches which centre thriving, resilience, resistance and joy (e.g., [35,36]). While this serves to mitigate perpetuating vulnerability and inferiority narratives which are discriminatory and prejudicial, our statements support calls for research and practice to go further to explicitly emphasise how social forces of privilege (both historic and contemporary) actively create, maintain and perpetuate inequities; on the ways in which advantaged groups keep others in disadvantaged positions and maintain the status quo [18,19,37–39]; and, to take a social justice perspective [40]. Our statements suggest that such an approach should also be incorporated into professional training and development in areas relating to social determinants of health.

### Community engagement, involvement and peer research

One definition of community engagement is “a range of approaches to maximise the involvement of local communities in local initiatives to improve their health and wellbeing and reduce health inequalities. This includes: needs assessment, community development, planning, design, development, delivery and evaluation.” [41], p.13). The importance of community engagement and involvement is increasingly acknowledged, operating on a continuum between consultative (organisation-led) and empowerment (community-led) approaches (e.g., [20]). However, without adequate resource and reflexivity they can risk being tokenistic, reinforcing existing power dynamics, creating burnout, and perpetuating extractive and exploitative relations (e.g., [24, 42]. The statements on these topics were supplemented by open text comments that highlighted how ‘good practice’ in community engagement initiatives require adequate time, training, resources, trust, explicit attention to power dynamics, and honest reflection on how health, care and research institutions drive health inequities through processes such as racism and (neo)colonialism (e.g., [24, 43–46]). Our statements also support calls for decision-makers, researchers and care providers to be supported and trained to take up findings from community-led work and/or to initiate good quality engagement activities [47]. Funders were also called on to take into account of what is needed to support researchers and communities in terms of time, resource and flexibility in order to avoid constraining engagement activities and perpetuating harm [48].

### Organisational culture change

Finally, two of our statements relate to greater equity within organisations in terms of both workforce (e.g., career progression) and working environment (e.g., related to Equity, Diversity and Inclusion initiatives). Diversity (e.g., in relation to race and ethnicity) within the caring professions has been linked to reduced inequalities for those being cared for (e.g., through greater concordance, improved access and uptake, greater cultural sensitivity)[49]. Inclusion of minoritised groups within the research community is similarly proposed to help produce research which more effectively addresses health inequities, for instance, via greater understanding of the needs and challenges of marginalised groups through shared life experiences, enhancing recruitment and trust in research, adoption of appropriate research methodologies, and development and implementation of interventions which best meet their needs [50–52]. Given the engrained and ubiquitous nature of organisational culture operating across multiple levels (e.g., shared assumptions, norms, values and beliefs), such culture change initiatives are difficult to implement [53,54]. Our statements emphasise the importance of attending to organisational culture including transparent monitoring of sustained and accountable mechanisms for identifying and enacting change, if efforts at recruiting, retaining, and promoting staff from minoritised groups within health, care, policy and research sectors are to be successful.

Areas in which consensus was not achieved may partly reflect the disciplinary backgrounds of respondents, the majority of whom were health and social care professionals and researchers which is likely to have shaped the wording, priority and nature of statements. For instance, two statements for which consensus was not achieved related to use of creative methods for research and dissemination. Respondents commented that they had a lack of experience with such approaches; however, they also suggested that while creative methods may be an important mechanism of capturing information shared by people underrepresented in research, they may not be perceived by decision-makers as sufficiently ‘robust’, hindering their impact. This suggests that a statement referring to understanding how and such evidence could be translated into decision-making may have been informative. There is evidence that arts-based interventions can work in subtle and complex ways to positively influence knowledge through emotions [55].

Further, open-text comments indicated that some of the non-prioritised statements: 1) should already reflect standard practice (e.g., relating to disseminating research in different formats, giving staff time and resources to ensure their work is fair and non-discriminatory), and/or 2) are not sufficiently impactful in themselves, and/or 3) were important but perceived as being downstream of other higher priority statements. Examples of the latter included statements relating to equitable career pipelines which are key but priority-wise would follow statements regarding amending organisational culture such that people are not being recruited into hostile or exclusionary environments. This could be seen as a circular argument in terms of how culture change might be achieved through better representation of those from excluded populations and highlights the complex interplay between institutional, community and individual dynamics [56]. The need to collect and share accurate data was perceived to be necessary for monitoring and equity impact assessments to be meaningful. Also, before sharing best practice, respondents highlighted that there is first a need to learn more about best practice from enacting and learning from other consensus statement-related activities.

Comments also suggested caution around statements involving guidance, frameworks, toolkits and so on, reflective of the fact that people can feel overwhelmed with ‘resources’. This may also point to the need for top-down action on a structural level to mitigate and/or counterbalance the need for individuals and organisations to take on additional work to address inequities. Finally, several comments reflected on the extent to which initiatives should be academic-led rather than community or service-led. While the statements are not meant to be read as academic endeavours, this perception may reflect the organisation of the HSE Collective, in which the Delphi process was organised and led by academic members, and which may have also influenced engagement with the survey more widely.

### Strengths and limitations

Our modified Delphi approach successfully gathered perspectives from a range of health and social care professionals, researchers with expertise across a range of health inequities topics, community organisation representatives, and a smaller number from policy and arts/culture backgrounds. Our approach differed from other prioritisation exercises in relation to health inequalities and public health which focus on agreeing research priorities (e.g., [57–59]). Instead, our statements aim to achieve consensus on ways of working which will help tackle the inherent structural and social forces within research, education and engagement/advocacy processes which serve to maintain and perpetuate inequities in place. The main limitation relates to the range of voices included. While our panel reflected diversity in relation to race, ethnicity, sexual orientation, migration status, health, and religion, the majority were educated to degree-level or higher and worked either as health and social care professionals or academic researchers. While we aimed to broaden the reach of the survey by asking people to share with their networks it is possible that the wording and approach (online survey) were exclusionary. Thus, future work involving sustained and in-person engagement to capture the perspectives of people and groups with lived experience of intersecting forms of social marginalisation (beyond those within research and health/social care fields), and voluntary and community sector organisation representatives is needed to extend and refine these priority ways of working. Another limitation was that only two statements met the 75% threshold, while the remaining nine were included as scoring high priority for over half of participants. We emphasise that future survey rounds incorporating a wider breadth of stakeholder representatives are needed to iteratively refine these principles to reach consensus and that this is also appropriate given shifts in social, funding and policy contexts.

### Conclusions

Health inequities persist and are widening despite sustained policy and research attention. The Health & Social Equity Collective’s prioritised statements offer a set of guiding principles for ways of working within research, education, and engagement/advocacy which would accelerate the pace of change in addressing health inequities and support the translation of research into practice. These principles complement initiatives which develop priorities for research topics in health and public health sphere, attending to how work is done. They have already informed the Collective’s approach in developing applications for research and education initiatives. Given the pressing need to address inequities, the principles provide a starting ground for future consensus building approaches which incorporate a wider diversity of perspectives, and to continually update these while integrating learning from health equity initiatives nationally and internationally such as the Accelerating City Equity Initiative [60]. Given the need to be contextually relevant and sensitive similar approaches could be taken in other geographic and social contexts to inform health inequalities practice.

## Data Availability

All data produced in the present work are contained in the manuscript

## Acknowledgements

We thank members of the Health and Social Equity Collective for supporting its co-design and for their involvement in creating these guiding principles, to maintain anonymity in this research we have not named Collective members individually. This paper represents independent research carried out by the Health and Social Equity Collective which received funding from the King’s Together initiative (King’s College London) and from Impact on Urban Health. S. L. H. and J.O. are part-funded by the NIHR Biomedical Research Centre at South London and Maudsley NHS Foundation Trust. C.W., and S.L.H. are supported by the ESRC Centre for Society and Mental Health at King’s College London (ESRC Reference: ES/S012567/1). The views expressed are those of the author(s) and not necessarily those of the funders.

